# Supporting Forced Migrant Fathers maternity journey in the UK; A mixed methods study for priority setting

**DOI:** 10.1101/2025.06.25.25330321

**Authors:** Andy Mprah, Melanie Haith-Cooper, Fiona Meddings

## Abstract

**Background:** Fathers, including migrant fathers, play a significant role in supporting maternal and child health. However, their experiences during the maternity journey are often overlooked, particularly in forced migrant populations. This study explores the barriers faced by forced migrant fathers during the maternity journey and identifies feasible interventions to improve their experience.

**Methods:** A two-phase qualitative study was conducted. Phase one utilized a virtual Nominal Group Technique (NGT) with six forced migrant fathers to identify and rank barriers to their maternity journey experiences. Phase two involved semi-structured interviews with six stakeholders, including midwives and voluntary sector workers, to assess the feasibility of interventions proposed in phase one. Data were analysed using framework analysis.

**Results:** The top three barriers identified by forced migrant fathers were: lack of community support centres, fear of being charged for maternity services, and lack of access to information about the maternity process in the UK. Suggested interventions included establishing community support centres, providing financial assistance, and increasing access to maternity-related information through simplified resources and mobile applications. Stakeholders highlighted the need for practical, cost-effective solutions such as extending existing services for mothers to fathers and providing staff training on forced migrants’ financial entitlements.

**Conclusions:** Addressing the barriers faced by forced migrant fathers requires a multi-sectoral approach, including community-based support, policy advocacy, and improved communication strategies. These interventions can enhance their involvement in the maternity journey, ultimately improving maternal and child health outcomes.

## Introduction

Fathers play a crucial role in maternal and child health, yet their experiences and challenges during the maternity journey remain underexplored, particularly among forced migrant populations (1). The transition to fatherhood can be complex, with factors such as cultural differences, financial constraints, and difficulties navigating the healthcare system adding additional layers of difficulty for forced migrant fathers (2). Research has demonstrated that paternal involvement is associated with improved maternal well-being, enhanced birth outcomes, and stronger parent-infant bonding (3). However, healthcare services and maternity policies often fail to fully integrate fathers into maternity care, leading to feelings of exclusion, distress, and limited engagement with healthcare services (4). These challenges are exacerbated for fathers who have been forced to leave their home country, who may face additional socio-legal barriers, language difficulties, and uncertainty about their rights and entitlements in host countries (5).

A previous study focused specifically on the maternity experiences of forced migrant fathers living in the UK (Mprah et al, under review). The study found that men faced numerous challenges, peculiar to forced migrant fathers, when supporting their pregnant partner through their maternity journey. Fathers reported cultural and language barriers, but also expressed fears around being charged for services which led to them feeling marginalised in the healthcare system. Tailored interventions are needed to address forced migrant fathers’ needs when experiencing the maternity journey, however, these interventions need to be prioritised. This paper reports on a study with the aim to identify the prevailing barriers experienced by forced migrant fathers during the maternity journey in the UK and to assess the feasibility of interventions to address these challenges.

## Materials and Methods

### Study design

A two-stage mixed methods approach was undertaken. Stage one involved a virtual nominal group technique (NGT) with six forced migrant fathers. The objective was for fathers to rank the order of importance of barriers experienced during their maternity journey with their partners in the UK and how they could be addressed through interventions. Stage two involved virtual semi-structured interviews with six key stakeholders. The objective was for stakeholders to assess the feasibility of developing and implementing the interventions in the real world. The study was approved through the Chair of the Health Sciences and Social Sciences Ethics Panel, University of Bradford, 18^th^ June 2020, reference E872.

### Stage one

#### Recruitment of participants

For stage one of this study, it was important that men could read and write, as well as speak English. Men who fitted this criterion and participated in a previous study exploring their experiences of the maternity journey in the UK (Mprah et al, under review), were invited to take part in this study. They had previously been recruited through voluntary sector organisations that provide support to perinatal forced migrant families. Five men from the previous study participated in the NGT. Four more men who had not been involved in the previous study were approached in the same way. Only one of these newly recruited men attended the NGT. Therefore, a total of six forced migrant men took part in this study. They had all experienced the maternity journey in the UK and were aged over 18. They lived in different areas of the UK and had therefore experienced different NHS maternity services on their journey.

Despite the optimum number of participants for a NGT exercise being 5-9 (6), having a smaller group made it more manageable with undertaking the NGT online and with men who had English as a second language. Other stakeholders who are commonly invited to participate in NGTs (6) were omitted to ensure the men felt able to speak out in a group context. Therefore, we studied stakeholders’ perceptions in part two of the study.

An information sheet was shared and discussed via WhatsApp with participants five days prior to the NGT event. Verbal consent to participate was then acquired. One hour before the event started, each participant was called by telephone to again discuss the study.

At the beginning of the event, the entire process was explained again to the participants and the right to withdraw. Verbal consent was obtained and audio recorded. Verbal consent is a well-established method which has been previously been used with this population, when there could be a fear of how a signature may be shared with the Home Office (7). All data were stored securely with an anonymous code in a locked cabinet at the office and on a password protected computer. Only the main author, AM, had access to the data. Pseudonyms were given to each participant to protect their anonymity in any written reports of the study.

Due to this group of men being potentially vulnerable, the researcher was aware that men may appear stressed during the NGT process, recalling difficult experiences. Mitigations were in place for this, stopping the process, using a breakout room to debrief the participant and providing follow up counselling if required

#### Procedure

Prior to the NGT, AM had previously consulted with a group of five forced migrant fathers who had participated in a previous study, exploring their lived experience of their maternity journey in the UK (Mprah et al, under review). Their role was to translate the themes/ sub themes from the previous study (see table 1) into six statements for ranking in this study, ensuring the wording was appropriate and the original meaning was not lost.

**Table 1.** Barrier statements co-produced from the themes from previous study (Mprah et al, under review)

| THEME | BARRIER STATEMENTS |
| --- | --- |
| Playing second fiddle | • Being side-lined at maternity appointments |
| Adapting to alien NHS maternity service | • Not able to take control of conversations because of language barrier and lack of interpreters.<br>• Lack of access to information about the maternity process in the UK |
| Living in the asylum system | • Lack of community support centres to support migrant fathers experiencing UK maternity.<br>• Fear of being charged and financial burden on the father as a breadwinner.<br>• Starting maternity appointments all over again due to dispersal to new location in the asylum process |

For this study the NGT process was undertaken (see Figure 1), audio recorded and transcribed verbatim. Firstly, using the six statements, the men individually ranked the statements from 1-6 in order of the most challenging barriers for them on their maternity journey with their partner in the UK. The men were given time to silently record their initial thoughts and ranking of the barrier statements. This was followed by the first of a series of four round robin discussions. The first-round robin involved participants individually talking about their rankings and the rationale for the order of importance to them. This was recorded on the Zoom whiteboard. After the group discussion, men were invited to modify their rankings if they would like to.

**Figure 1.**
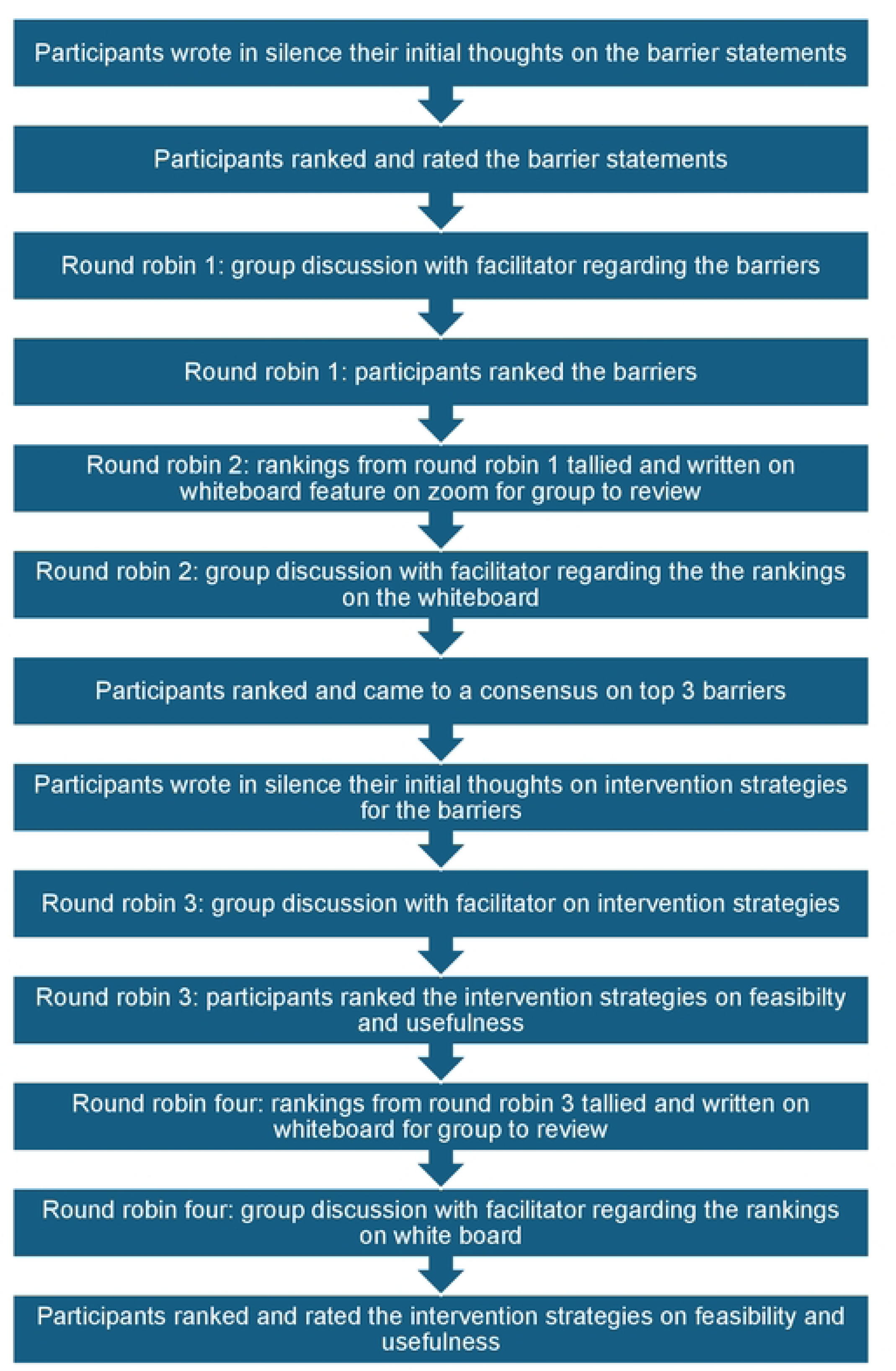
The Nominal Group Technique Ranking Process.

In the second-round robin discussion, each participant’s ranking of the barrier statements was re-recorded on the whiteboard. Participants who changed their minds about the ranking were invited to discuss their change with the group. The rankings from the first-round robin discussion were compared with the second in a group discussion. Participants then came to a group consensus on the top three statements which they felt reflected the barriers forced migrant men experienced on their maternity journey with their partners in the UK. Each participant was then asked to write in silence their initial ideas of interventions that might address the top three barriers.

The third-round robin involved a group discussion about the participants’ ideas for interventions. This was recorded on the whiteboard and the participants individually ranked each intervention, in the context of feasibility and usefulness. In the fourth-round robin, individual rankings of the interventions were tallied and recorded on the whiteboard. A group discussion was then facilitated to reach a group consensus on how the interventions ranked in terms of feasibility and usefulness.

### Stage two

#### Recruitment

Six stakeholders were purposefully selected based on their firsthand knowledge and experience of working with forced migrant families experiencing the maternity journey (see table 2). Four stakeholders had backgrounds in the voluntary sector (one a midwife) and two were academic midwives involved in research in this area. We aimed to include stakeholders working in the local authority in the study but due to the COVID-19 pandemic, they were unable to commit the time to an interview. The six participants were based in five different areas of England. Information sheets were e-mailed to them and any queries were addressed. Verbal consent and the right to withdraw were audio recorded at the beginning of the Zoom interview.

**Table 2.** Stakeholders’ demographics.

| Pseudonym | Designation |
| --- | --- |
| Claire | Midwife |
| Kate | Midwife in the voluntary sector |
| Fatia | Voluntary sector officer |
| Sue | Voluntary sector officer |
| Anne | Voluntary sector officer |
| Alice | Midwife |

### Data collection

Semi-structured interviews were conducted via Zoom with the six stakeholders to discuss the forced migrant fathers’ barriers on their maternity journey in the UK with their partner, their proposed interventions and the feasibility of implementing these in the real world. The interviews were audio recorded and transcribed verbatim.

### Qualitative data analysis

The qualitative data set for both phase one and phase two of the study were analysed together using framework analysis (8). Transcripts from the nominal group technique and the individual qualitative interviews were uploaded onto NVivo to organise the data. AM undertook a process of familiarization; reading and re-reading the transcripts whilst listening to the audio recordings and noting any a priori and emerging issues (see table 3). MC and FM independently cross-checked the emerging themes against the transcripts to ensure this process was robust. Stage two involved developing framework categories from the a priori and emergent issues to organise the data, then indexing the data by revisiting each transcript and extracting the preliminary codes into the framework categories.

**Table 3.** A priori and emergent themes.

| **A priori themes** | **Emergent themes** |
| --- | --- |
| Letters addressing payments | Using children's centres |
| Online platforms | Parent educator midwives to communities |
| Staff training | Safe care for fathers |
| Copying existing models | Continuity of carer |
| Virtual platforms | Certificate of exemption |
| Posters with maternity pathway | Diagrammatic information in statutory documents |
| Mobile applications | Activism on payments |
| Improve hospital information | Payment plan |
| Establish more community support centres | Different hubs for fathers within existing programs |
| Hotline for information | Brochures to show responsible clinician |
| Leaflets | Empower voluntary sector |
|  | Sensitivity in gender of midwives |
|  | Health providers to lead discussion in community |
|  | Staff keeping up to date |
|  | Building on what exists |
|  | Health visitor visits |
|  | Update statutory maternity documents |

After indexing each interview transcript, AM used NVivo software to extract all data coded to a framework category for a particular participant. The framework categories from the stakeholders either supported or conflicted with the forced migrant fathers’ framework categories. 28 preliminary codes were grouped together to illuminate the most important aspects of the data related to the research question. To ensure the framework categories were based around the issues arising from the data and to refine the process, we applied the framework to two initial transcripts. Next, we undertook the charting stage to organise the data into a more manageable format. This involved summarising the data in each framework category which was then linked to the relevant text in each transcript in NVivo. Finally, the data were mapped and interpreted. This involved using the charted data to find patterns in the data, then developing themes to capture the nature of the interventions from the men’s and stakeholders’ perspectives.

These preliminary a priori and emergent themes (table 3) were organised in NVivo as framework matrices to develop a static model for all themes (see Figure 2).

**Figure 2.** Original Static Model of initial themes in NVivo 12.

#### Findings

The findings from the ranking exercise and the respective final rankings of the barrier statements from the forced migrant fathers are shown in tables 4 and 5.

**Table 4.** Ranking collation findings.

| **Pseudonyms** | **Zizou** | **Raman** | **Nitin** | **Saad** | **Ahmed** | **Ishak** | **Total** |
| --- | --- | --- | --- | --- | --- | --- | --- |
| **Statements** |  |  |  |  |  |  |  |
| Being side-lined | 4 | 6 | 1 | 1 | 2 (6) * | 5 | 19 (4<sup>TH</sup>) |
| Fear of being charged and financial burden on the father as a breadwinner | 2 | 2 | 6 | 3 | 6 | 2 | 21 (2<sup>ND</sup>) |
| Not able to take control of conversations because of language barrier and lack of interpreters | 1 (5) * | 5 | 5 | 2 | 3 | 3 | 19 (4<sup>TH</sup>) |
| Lack of access to information about the maternity process in the UK | 3 | 4 | 4 | 4 | 1 | 4 | 20 (3<sup>RD</sup>) |
| Starting maternity appointments all over again due to dispersal to new location in the asylum process | 6 | 1 | 3 | 5 | 1 | 1 | 17 (6<sup>TH</sup>) |
| Lack of community support centres to support migrant fathers experiencing UK maternity | 6 | 3 | 2 | 6 | 2 | 6 | 25 (1<sup>ST</sup>) |

Key: \* Father after discussions among peers changed his mind about the ranking of a particular statement and gave a different ranking

**Table 5.** Final barrier statement ranking based on consensus.

| Barriers | Ranking |
| --- | --- |
| **Lack of community support centres to support migrant fathers' experiencing UK maternity** | (25) 1<sup>st</sup> |
| **Fear of being charged and financial burden as a breadwinner** | (21) 2<sup>nd</sup> |
| **Lack of access to information about the maternity process in the UK** | (20) 3<sup>rd</sup> |
| **Being side-lined at maternity appointments** | (19) 4<sup>th</sup> |
| **Not able to take control of conversations because of language barrier and lack of interpreters** | (19) 4<sup>th</sup> |
| **Starting maternity appointments all over again due to dispersal to new location in the asylum process** | (17) 6<sup>th</sup> |

The findings from both stages are reported together below based on the prevailing top three barriers in table 5. Table 6 presents the top three barrier statements

**Table 6.** Themes and sub-themes from the merged dataset.

| Themes | Sub-themes |
| --- | --- |
| **Community support centres.** | ❖ Multipurpose community support centres<br>❖ Dedicated support in the community from same health providers |
| **Providing financial support** | ❖ Maternity benefits<br>❖ Strategies to address charging.<br>❖ Staff training on financial entitlements<br>❖ Information for forced families on NHS entitlements |
| **Increasing maternity knowledge** | ❖ Maternity pathway diagrammatic illustrations<br>❖ Health providers-lead discussions in community<br>❖ Online/digital platforms |

### Community support centres

Three of the forced migrant fathers ranked a lack of community support as the most challenging barrier to their experience of the shared maternity journey in the UK and gave it a score of six. One of the fathers compared his experience of community support with a previous pregnancy back home:

> *“Not having support from the community compared to back home during my wife’s condition was our biggest challenge with my wife’s pregnancy in the UK and that is why I ranked this as the biggest barrier for me.” (Saad)*

This comparison with back home was reflected in the stakeholder’s discussion:

> *“I think the first barrier sort of embodies socio-cultural differences between the UK and back home, hence the fathers asking for the support which they would have gotten back home.” (Sue)*

To address this barrier, all fathers shared their views about establishing multi-purpose community support centres in their communities for migrants as a ‘one-stop shop’ to provide support to fathers when their wives are pregnant. One father stressed the importance of interpreters being available in these centres for the most commonly spoken languages:

> *“A lot of people struggle with the English language and having a lot of interpreters especially in the Arabic language will be more helpful. Most of the migrants speak Arabic and lack proficiency in the English language. I know it will be impossible at these centres to have interpreters for all languages of the world.” (Raman)*

Another father reported on how this could address the difficulties experienced with hospital interpreters by having local interpreters in the community settings:

> *“We are Arabic speakers, but there are different dialects with Arabic, so my wife and I struggled with some words from hospital interpreter. He spoke a different dialect of Arabic. If there was a community centre, I would find someone who speaks my kind of Arabic.” (Ishak)*

Stakeholders’ responses to the suggestion of community centres for fathers varied. Two stakeholders questioned whether men would attend such a centre due to cultural beliefs about pregnancy and childbirth:

> *“Well, it is an interesting one. I can hear them say it, but the question is will they attend it? I ran a group for people seeking asylum and for refugees. But trying to get men attend itself is a barrier (laughs). There is this cultural thing that makes them question if maternity is a man’s business.” (Anne)*

All the stakeholders agreed that adding to existing community support services would be easier and more efficient to establish than creating new services:

> *“If you talk about cost-effective interventions, what already exists for women can be branched to have same for the men. So rather than starting from zero for the men, we can have an extension of the services already available to the migrant women for the migrant fathers. It will be easy to say bring the fathers along and this can be a way of making the fathers meet.” (Fatia)*

This could be through providing space in existing accommodation, including children’s centres, but also other places providing antenatal care:

> *“To have a breakout session within the antenatal classes for the forced migrant fathers to meet with other fathers. By this the husband knows the wife is in a safe place and he can also have information about maternity tailored to his needs.” (Alice)*

One stakeholder also suggested copying a nationally known model of support for men:

> *“There are groups around like men’s club which provides support to men and has been a success nationally. I wonder if the success from that model can be learnt and practiced with migrant fathers.” (Sue)*

Another stakeholder suggested that it would be challenging for midwives in the UK, who are predominantly white, to talk to forced migrant fathers at the community support centres about how to navigate the maternity journey as it may be from the wrong perspective. She was in support of this intervention but thought it should be run by the fathers:

> *“It is a clever idea, but it should be run by the right people. It will be great to kind of train the migrant fathers to kind of manage the community centre and then have midwives there to answer the maternity questions.” (Alice)*

Further to this, another stakeholder suggested that fathers could provide peer support to one another in the community centres. However, another stakeholder felt that a peer support model would not work due to cultural beliefs about the role of the man:

> *“Just that men do not seem to want peer support because they feel they are men. Exactly, most of the men do not want to appear to be seeking for help.” (Fatia)*

Most fathers suggested that seeing the same health professionals at the community support centres during the maternity journey would make them feel more confident. Stakeholders agreed with the men that the same midwives and other health providers should access migrant fathers within the community environment to lead discussions. They suggested that continuity of carer where the same midwife continues to see migrant families, would increase trust and confidence, and help increase knowledge of maternity processes:

> *“I think that for this we need to go down the continuity of carer route. That means if there are more scans, appointments, and tests with consultants…if they have continuity of carer midwife, then the midwife will be able to guide them. This will make the migrant fathers more involved and not coil in their shells.” (Sue)*

One stakeholder stressed the importance of health professionals seeking out the fathers rather than expecting the fathers to find them:

> *“You are looking at this, the health providers do want to engage men in different sort of ways and the answer could be going around the group in a community centre rather than expecting them to come to you. What makes a huge difference from my experience is people having space to talk through their fears and experiences and you do not experience that without going to them.” (Anne)*

However, another stakeholder suggested that health professionals need to reach out to migrant fathers and their partners in their homes instead of the community centres:

> *“It might be difficult to get around some of these centres and might need to do more home-based visits to the migrant families. Midwives need to be able to individualise the maternity services to the forced migrant families by going to them.” (Claire)*

### Providing financial support

The financial burden of being a forced migrant father was ranked as the second most challenging barrier, with two fathers ranking this barrier with the highest score of six. Both fathers and stakeholders discussed how the fathers’ financial circumstances in the UK were different to back home where they were working and financially independent:

> *“I have seen fear. Certainly, the financial burden on the father as a breadwinner is massive and boils down to being able to feed your pregnant wife and to attend maternity appointments which he cannot because he has no money.” (Sue)*

To address the financial burden, all the fathers suggested a form of financial support, financial waiver, or vouchers should be provided to assist forced migrant families during the maternity journey in the UK. One stakeholder agreed that providing some form of monetary assistance for forced migrant families expecting a baby would help them in their maternity journey:

> *“I think what they are saying is for them to be provided some form of maternity benefits. This is something that could be done easily without costing the Government so much. I think they should be an exception and given all the maternity benefits they require.” (Alice)*

Being charged for maternity care was a big fear for men, with one father discussing his specific situation around being charged when he should not have been:

> *“I still received a bill from the NHS of £13,286. And it was stressful for me. This has happened not only to me. It has happened to lots of people. Because of the pressure I went through, I chose this barrier as the biggest and first for me.” (Nitin)*

Five of the stakeholders highlighted how bills for maternity care are sent to women rather than their partners, reflecting the maternity model which focuses on the woman. This they believed, increased fear in the father who were not given any information about charging. The fathers all agreed that they need more information about charging for maternity care through organised events:

> *“Also, the Home Office should make it like errm, for example we have drop-in sessions for the asylum seekers and refugees one day a week. So, they can use this to explain to them what these charges are about. Because I used to volunteer for these kinds of things about being charged and advising people that if you go to the GP and you do not tick the right box, you might be charged or sent a bill.” (Ishak)*

Some stakeholders agreed that this was feasible and suggested the need for such information to be included as a package for forced migrant families at their point of entry into the UK. However, this information should come from the NHS rather than the Home Office. One stakeholder reported that:

> *“I also think there should be like a central package of information with different information to be given to forced migrants on arrival. This could include how to register with a GP, where to start if your partner is pregnant and the stages involved for both normal and abnormal pregnancies, how to access a midwife.” (Kate)*

One father suggested that the NHS should automatically recognise the fact that both refugees and asylum seekers are displaced people in the UK and that any financial obligation with maternity services is waived completely:

> *“The people at the hospital should know we are here and have nobody and no money and so payment for my wife to have a baby should not be an option at all” (Ahmed).*

This could be recorded in the maternity notes:

> *“Maternity notes and books should contain information about our eligibility for free maternity services, so that the midwives can understand what we are entitled to without asking us if we are eligible.” (Saad)*

Stakeholders agreed with this idea, one specifically discussing how this could work in practice:

> *“I think a national maternity note with a page added to address charging issues and eligibility for forced migrants is doable. The notes can also be online, and the contents can be interpreted using google translator into the local languages of the respective migrant fathers. And because it is a national document, they will not face the ordeal of being billed for maternity care.” (Claire)*

Some of the stakeholders supported the idea of a certificate or letter to prove eligibility for free NHS maternity care. One stakeholder reported:

> *“Yeah, it is feasible and if it comes in bowth ways. So, the information provided in leaflets and certificates to be given to the family. Something the migrant family can hold on to so that they can be empowered to produce when they are asked to pay for maternity services.” (Sue)*

However, other stakeholders were not as keen on statutory documents capturing issues about maternity charges. One stakeholder cited the ever-changing policies on immigration and felt charging needed to stop altogether:

> *“I think the issue is that such guidance changes rapidly to be put in a statutory document. I think charging should be stopped full stop. The economic cost of generating bills that terrify families is far higher than any thumb they claw back in terms of payment.” (Alice)*

Another stakeholder reported a strategy asylum seeker fathers could use to deal with their partners being billed for maternity services:

> *“In fact, what we have done is to work with the overseas charging department where we wrote a letter that if migrants are chargeable, we get them to adapt this letter to send to the overseas department. And, it says, I have received a bill, am homeless. I would like you to consider the bill and if not, I would like to make a payment plan of a penny a month and I would need to pay this in cash because I do not have a bank account…Now because they have made a payment plan, and sends that in, the trusts are writing off the bill because they do not accept cash payments.” (Anne)*

Stakeholders and migrant fathers agreed that maternity staff and senior staff in the NHS should be updated on the entitlements to free maternity care of forced migrant families through staff training. This could be through the annual staff training already set up to maintain competency:

> *I think there should be annual or regular training for staff to be updated on this. Because if from the ranking it is coming out as number 2, as the second most important barrier to maternity then it means lots of people here have bad experiences with the charging”. (Raman)*

One stakeholder identified that this approach would be cost effective:

> *“There is a yearly training for staff about safe cares. The issue about migrants can be included in yearly updates and that will not cost any money because it happens in the UK anyway.” (Sue)*

Another stakeholder identified that national guidelines for maternity care of forced migrant families also need updating:

> *“If you look at the NICE guidance for the care of people with complex social histories which includes forced migrant families, it does not say very much about how to care for such families well. But one of the few things it does say in the guidance is that midwives should be aware of procedures for who is eligible to be charged for treatment.” (Alice)*

### Increase Knowledge of NHS maternity processes

A lack of information about NHS maternity care was ranked as the third most challenging barrier influencing the forced migrant fathers’ experience of

their maternity journey. Although none of the fathers gave this the highest score, four fathers gave it a score of four. These fathers compared the journey to back home where they had adequate knowledge of the maternity process:

*“I was always confused what next, who we must see and where we must go, when my wife was pregnant compared to Pakistan. The midwife treated it like we already know it all and that is why I ranked this as my third biggest barrier” (Raman)*.

The stakeholders all agreed there were gaps in information about the maternity processes in the UK among migrants, especially for forced migrant fathers and the need for targeted interventions to address this. One Intervention suggested by the fathers was simple charts and diagrams clearly illustrating the NHS maternity services journey in the UK.

*“My English is limited so a simple chart in the maternity note showing me where to go next and who to see next with our pregnancy will make me confident to accompany my wife to appointments.” (Ahmed)*

Stakeholders agreed with this suggestion, developing such diagrams in the maternity notes:

> *“Something like an info gram with diagrams and not something wordy. So instead of the national platform, the simplified information in the antenatal notes where one does not need to be a speaker of English to at least know what will happen. I think doing things like these to enable the migrant fathers to effectively assist their partners and be there for them will go a long way to improve maternity outcomes of the women.” (Claire)*

Another stakeholder discussed what may be included in the diagrams:

> *“Like what to expect when labour happens, and things to improve the confidence. Things about health visits and what will happen at every point in time including scans, HIV tests as well as who else will be involved in the maternity care. I think this could certainly be able to go into the antenatal notes.” (Sue)*

However, another stakeholder suggested such simplified illustrations in maternity documents could be used as a tool for the midwife to discuss the maternity process and avoid unnecessary confusion:

> *“And if such diagrams are produced in the maternity notes or red book, the midwives will be able to use the diagram as an additional resource to take them through and explain to them what to expect at each stage of maternity. I support this approach rather than the diagram being produced in the red book to be the answer.” (Sue)*

Most fathers and stakeholders agreed that mobile applications (Apps) could be helpful to increase their knowledge of the maternity journey. Apps could be safe spaces for fathers to seek information about important stages of their partners’ pregnancy. One father reported that this would help fill the gap in his understanding due to not being able to attend appointments with his partner:

> *“I work at night in a fruit factory and when I come home, I get tired and sleep all day. I was not able to attend any meeting about my wife’s pregnancy. May be if there was an app where I can follow and learn everything about the pregnancy, that would be very good for me.” (Razak)*

Stakeholders suggested that it would be more efficient and cost effective to adapt existing mobile apps for forced migrant fathers rather than building one from scratch. They also discussed the importance of a mobile app including features where the father is signposted to appropriate professionals, should they need further advice:

> *“Maybe there should be a search tool there to signpost migrant fathers to the right information about their wives’ pregnancy. So, a link within the app or a helpline… I think something like that will sit well within existing charitable organisations.” (Claire)*

However, another stakeholder raised concerns about the management and updating of the app and also issues around confidentiality if the father had to create an account. Other stakeholders suggested that the app be controlled by the fathers themselves, to ensure it meets their needs:

> *I do not think that we should control them. They are so isolated in the community. I think people in the health sector try to control and regulate things and all these forced migrant fathers have endured control over their lives. Actually, it feels more like this needs to be led by them.” (Alice)*

There were however practical concerns raised around fathers using the app including their digital literacy levels and also the cost implications:

> *“They (the fathers) must be digitally included, and I do not think that all of them have the money for the internet. So, I think that will exclude people let alone the language and ability to use it.” (Anne)*

## Discussion

The aim of this study was to identify the most challenging barriers experienced by forced migrant fathers during the maternity journey in the UK and to assess the feasibility of interventions to address these challenges. The top barrier was a lack of community support which fathers thought could be addressed by setting up community support centres for men with dedicated support from health professionals. The second barrier was the financial burden of being a forced migrant father and breadwinner. The men believed more education was needed about charging for maternity care targeting forced migrant families and NHS staff. Stakeholders supported this, and also providing extra benefits for asylum seeking families accessing maternity care. The third barrier was lack of access to information about the maternity process in the UK, which could be addressed through simple charts and diagrams and mobile phone applications (apps) to inform fathers about the UK maternity journey. A pattern across all three themes was the stakeholders’ suggestions that to be cost-effective, adapting current resources including community services, maternity notes and mobile apps to meet the needs of forced migrant rather than creating something new.

The fathers suggested establishing community support centres in areas where forced migrant families are predominantly living. They identified socio-cultural differences to back home where communities of friends and family around them supported their maternity journey, reflecting a previous Canadian study (9). However, these centres need to be established in a culturally appropriate way to address the socio-cultural needs of migrant families. Reflecting previous research with Somali refugee fathers (10), involving men in co-producing these services would ensure their socio-cultural needs are met.

There is a paucity of research around the practicalities of establishing community support centres in a maternity context for this population. However, a systematic review reported how accessible health clinics in terms of location and opening times, facilitated refugees attendance for care (11). More recently, a Turkish study of Syrian refugees found that centres located in the local refugee communities were effective in improving uptake of health services and health outcomes (12). Similar to our findings, (13) found that existing buildings including schools could be converted into multifunctional community centres to support and integrate refugees and migrants.

The financial burden was another challenge for fathers experiencing the maternity journey in the UK. They came from countries where they were financially independent and were now living in poverty and fearing being charged for maternity care. Both fathers and stakeholders believed that more financial support was needed for forced migrant families and there should be no charging for services. A recent scoping review, linked the fear of being charged for health services with a decline in mental health (14). It would appear counterproductive to charge for care when people cannot afford to pay whilst exacerbating poor mental health which could deter people accessing timely care (15) leading to a requirement for increased health care. A lack of consistency and inappropriate charging for services, and a need for an exemption certificate was raised by both fathers and stakeholders, reflecting previous research (16).

The need for more Information and education was raised across the themes, related to NHS staff requiring training around charging for services and the wider sociocultural dynamics and patriarchy of forced migrant families. Most of the fathers suggested that back home, they were knowledgeable about the maternity journey, and this made them confident as fathers. Here, the fathers needed more information around charging, and the wider maternity journey. Previous literature has focused on further training of health professionals to meeting the needs of forced migrant women (17) but not included the needs of fathers in this training. Similarly, the literature discusses providing information packages, posters, leaflets and drawings (18)and drop-in sessions for migrant women (19) but not fathers.

These is a body of literature around the use of apps and other digital/virtual platforms to provide information for forced migrant families (20, 21). A study with Syrian refugees discussed the importance of considering families health beliefs, literacy levels and the family structure when developing digital technologies to support access to maternity care (21). The DAISI project (22) used behaviour change theory to co-produce and feasibility test a digital animation intervention in two languages, to enhance postnatal women’s awareness of sepsis and how to reduce their risk of developing the condition. It was particularly useful for migrant women and was considered acceptable, culturally sensitive, and simple to implement and follow. Initiatives such as the DAISI project have the potential to increase information giving about the maternity journey more widely with diverse groups. Forced migrant fathers could benefit from this type of animation, being available on smart phones.

The findings suggested that there was a mismatch between the perspectives of the fathers and stakeholders around the forced migrant father’s involvement in maternity care. For example, some stakeholders believed that fathers would not attend community support centres if they were set up, in contrast to the fathers who requested these as an intervention. The fathers originated from different areas of the world. They reported cultural differences in childbirth between their country of origin and the UK. In some areas, men reported that childbirth was culturally seen as ‘women’s business’ and men were not involved in pregnancy and childbirth. In contrast, in other areas men reported being in charge of childbirth, paying the male doctors to care for their wives. All men discussed how they had to adapt to a new role in the UK, either becoming involved in childbirth due to a lack of support from female family, or relinquishing control, due to the UK cultural norm being women making decisions about their care. To take on this cultural norm in the UK, forced migrant fathers reported needing information about the maternity journey. The mismatch between stakeholders and men’s perspectives suggests more education is needed for stakeholders around forced migrant fathers’ inclusive role in the UK in order that services and interventions meet their perceived needs.

### Strengths and limitations

This study provided a platform for stakeholders and forced migrant fathers to be involved in the same research around the maternity journey experience of these men. However, they were engaged separately to ensure the voices of the forced migrant fathers were heard. However, this may have also negatively influenced the quality of the discussion in the NGT, where a dialogue between all members triggers the ranking and consensus of the statements (6). However, this study ensured all men felt able to contribute by removing any perceived power from other stakeholders in the group. One of the men involved in this study had not been involved in the previous research that led to the barrier statements for ranking. This may have led to this man’s not understanding the meaning or relevance of the barrier statements presented to him, influencing the quality of his input. Also, not including non-English speaking men, who may experience enhanced communication barriers may have excluded a particular narrative around language barriers in maternity care.

## Conclusion

This study has presented and discussed the findings from a NGT ranking exercise followed by qualitative interviews with key stakeholders which facilitated the gathering of a data set around forced migrant fathers’ three prevailing barriers to the shared maternity journey in the UK. The findings of both phases of this study were discussed and the need for co-produced interventions, building on existing services to meet the needs of forced migrant fathers.

By exploring both migrant fathers’ perspectives and stakeholder insights, the study sought to generate evidence-based recommendations for improving paternal engagement and support in maternity care, ultimately contributing to better maternal, paternal, and infant health outcomes.

## Data Availability

Data cannot be shared publicly because of protection of the participants who are forced migrants. Data are available from the University of Bradford Institutional Data Access / Ethics Committee (contact via authors) for researchers who meet the criteria for access to confidential data.

## Declarations

### Ethical approval and consent to participate

Ethical approval was granted by the Health and Social Sciences Ethics Panel of the Faculty of Health Studies, University of Bradford for this study with Ethics Application reference E775. Due to the COVID-19 pandemic and the nationwide lockdown in the UK, further approval was sought to adopt virtual semi-structured interviews instead of a face-to-face one. An approval email from the Chair of the Research Ethics Panel of the University of Bradford was received on June 18, 2020, for the changes requested. This primary study was conducted in accordance with the Declaration of Helsinki.

### Human Ethics and Consent to Participate

All forced migrant fathers in this study were taken through and issued an appropriate informed consent form to sign before participating in this study without coercion.

### Consent for Publication

Not applicable.

### Availability of data and materials

The datasets generated or analysed during the current study are not publicly available due to this review being part of a PhD research. They are available from corresponding author upon reasonable request.

## Acknowledgements

We acknowledge all voluntary sector organisations, forced migrants and other stakeholders who assisted in this study.

## Ethics approval and consent to participate

Ethical approval was granted from the University of Bradford Ethics Committee reference E775 on June 18, 2020. Participants gave verbal informed consent audio recorded and transcribed as evidence.

